# Community ECHO (Extension for Community Healthcare Outcomes) Project Promotes Cross-Sector Collaboration and Evidence-Based Trauma-Informed Care

**DOI:** 10.1101/2021.07.16.21260670

**Authors:** Christina A. Buysse, Barbara Bentley, Linda G. Baer, Heidi M. Feldman

## Abstract

**Background:** Adverse Childhood Experiences (ACEs) are traumatic events that occur before age 18 years. ACEs, associated with negative health behaviors and chronic health disorders, disproportionately impact people from poor and marginalized communities. Toxic stress from ACEs can be prevented and treated with trauma-informed care. Inadequate training prevents the maternal and child workforce from providing evidence-based trauma-informed care. Cross-sector collaboration between pediatric care sectors is crucial to providing systems-wide trauma-informed care, but significant barriers impede cross sector communication. Training and formal cross-sector communication networks are needed to create strong systems of trauma-informed care in communities.

The Stanford ACEs Aware ECHO (Extension for Community Healthcare Outcomes) program was created with 3 workforce development goals: 1) introduce the California Office of Surgeon General-led ACEs Aware Initiative to the maternal and child health workforce in 3 California counties, 2) disseminate trauma-informed evidence-based best practices, 3) bridge community silos to increase collaboration between care sectors to promote trauma-informed care systems.

**Methods:** Participants were recruited from Federally Qualified Health Centers, county public health departments, community behavioral health organizations, educational institutions, and agencies that serve low-income children and families. 100 unique participants representing 3 counties and 54 agencies joined sessions. Twelve virtual educational sessions were convened over 6 months using the Project ECHO® model via Zoom technology. Sessions consisted of didactic lectures and whole-group case-based discussions.

**Results:** After completing the educational series, participants expressed commitment to increased cross-sector collaboration, and reported increased knowledge and confidence in using trauma-informed skills. After participation, a significant number of participants had also completed another recommended California ACEs Aware Initiative online training.

**Conclusion:** An ECHO series of virtual workforce development sessions on trauma-informed best practices promoted cross-sector communication and was associated with strong participant engagement and satisfaction. The educational series increased knowledge and confidence in use of evidence-based trauma-informed best practices.

**Key Messages:**

- Adverse Childhood Experiences (ACEs) are stressful or traumatic events that occur before the age of 18 years and are associated with high human and economic costs to society.
- Evidence-based trauma-informed Care (TIC) has been shown to prevent and treat toxic stress caused by ACEs, but training programs for the maternal and child workforce have historically provided inadequate training in TIC best practices.
- Cross-sector collaboration to provide systems-level trauma-informed care is known to improve population health and promote health equity, but funding and staffing barriers to optimized collaboration exist.
- The Project ECHO® (Extension for Community Healthcare Outcomes) model uses distance learning teleconferencing to create learning communities across geographic distance and care sector, educate and mentor participants, and elevate quality of care in community settings.
- The Stanford ACEs Aware ECHO effectively delivered a curriculum about TIC to a tri-county community of maternal and child workforce participants that promoted cross-sector collaboration. Participation was associated with high levels of engagement and satisfaction. Participants reported increased confidence and knowledge to provide systemic evidence-based TIC.

## Introduction

Adverse Childhood Experiences (ACEs) are stressful or traumatic events that occur before the age 18 years. When first described in the Centers for Disease Control and Prevention (CDC) and Kaiser Permanente Adverse Childhood Experiences Study in 1998, 10 specific ACEs in childhood were described: physical, emotional, and sexual abuse; physical and emotional neglect; household challenges caused by a household member who experienced mental illness, used substances, experienced intimate partner violence, was absent due to divorce or separation, or was incarcerated.(Centers for Disease Control and Prevention, 2019a; Felitti et al., 1998) Exposure to ACEs has been associated with poor health outcomes, increased behavior risks, and decreased educational and economic outcomes. Adults who have experienced 4 or more ACEs are 1½ times more likely to have cardiac disease, 2 times more likely to have cancer, 5 times more likely to be depressed, and 10 times more likely to become substance-dependent than someone with no exposure to ACEs.(Bhusan et al., 2020) Children exposed to ACEs are at increased risk for behavioral, learning and attentional difficulties.(Jones et al., 2019) When accounting for direct healthcare costs and disability-adjusted life-years, it is estimated that ACEs cost $748 billion in North America. (Bellis et al., 2019) Societal costs of ACEs exceed hundreds of billions of dollars each year.(Centers for Disease Control and Prevention, 2019a)

Sixty percent of adults across many US states report at least 1 lifetime ACE, and 15.6% report 4 or more ACEs.(Merrick et al., 2018) The social determinants of health, such as poverty, systemic racial injustice, and housing or food insecurity exacerbate the negative impact of ACEs on health. It has been widely established that people who are racially marginalized, in lower income brackets, involved in the justice system, women, and members of the lesbian, gay, bisexual, transsexual, queer (LGBTQ+) community experience higher rates of ACE exposure than individuals without those characteristics in the general population.(Bhusan et al., 2020)

To prevent or mitigate the adverse effects of ACEs, the CDC recommends creation of programs in communities and schools that combine and amplify results to prevent violence, treat stress, and provide social and economic supports.(Centers for Disease Control and Prevention, 2019b) Public health leaders recognize that workforce development, a culture of collaboration, and systems strategies are necessary to bring about health equity.(Carlin & Peterman, 2019; Chandra et al., 2017) With a strong commitment to state-wide cross-sector collaboration and the bold goal of decreasing ACEs in half in one generation, the *blinded* Office of the Surgeon General and the Department of Healthcare Services launched the ACEs Aware Initiative in 2019. This multi-pronged public health initiative seeks to educate the maternal and child health workforce in the science and treatment of toxic stress, increase screening for ACEs, and promote wide-spread institution of trauma-informed care best practices in the state.(Bhusan et al., 2020)

Trauma-informed care (TIC) can be conceptualized as an overarching framework for providing care to individuals that experience ACEs and other trauma. TIC addresses the individual from a strengths-based perspective, with a goal of preventing and mitigating trauma at multiple levels of community and function. TIC requires understanding of the science of toxic stress and recognizes that trauma can be prevented and addressed at the primary, secondary, and tertiary level. At the individual level, trauma-informed care can be integrated into clinical encounters, parenting support, educational, and judicial systems. At the service systems level, effective programs can reach individuals who have experienced trauma and promote positive experiences through the healthcare, educational, child care, behavioral health, child welfare, criminal justice, and recreation systems.(Tebes et al., 2019) To be effective at the public health level, TIC requires a common language and functional communication network among providers, programs, and care sectors. TIC impacts all aspects of care delivery and requires effective communication between cross-sector care teams.

The maternal and child health medical, mental health, and direct service workforces are key to identifying and addressing the immediate health needs of individuals and families. Front line providers have a unique opportunity to intervene at all prevention levels. Barriers to implementation of systems of trauma-informed care have been identified across and between care sectors. Medical education training programs, for example, have historically not prepared primary care clinicians (PCCs) adequately to deliver trauma-informed community care. Clinicians in Pediatrics, Family Medicine, and Obstetrical care report that they lack adequate training and time to manage complex mental health problems and identification and care of trauma in their patients.(Dichter et al., 2018; Horowitz & Cousins, 2006; Horwitz et al., 2015) Mental health providers also report a lack of time and training as barriers to providing TIC. (Sharif et al., 2021) Gaps in trauma-informed care systems include lack of sustainable funding for providers to identify and treat toxic stress, lack of resources to mitigate secondary trauma for staff, and barriers to access to services once trauma exposure has been identified.(Bargeman et al. 2020)

Workforce development efforts have been shown to directly increase the capacity of community providers to screen and manage ACEs, mental health disorders, and adverse outcomes associated with social determinants of health in women and their children.(Kerns et al., 2016; Palfrey et al., 2019) A professional development session that provided trauma-informed educational strategies to educators in nursing and criminal justice fields led to significant increases in knowledge of trauma-informed education and to self-perceived increases in ability to support students impacted by trauma.(Doughty, 2020) Participants across sectors of education, law enforcement, judicial system, and mental health demonstrated increased familiarity with trauma-informed care when monthly cross-sector meetings were created.(Clements et al., 2020) PCCs who have received recent continuing medical education (CME) on mental health concerns screen more frequently for these issues than those who have not received the CME.(Green et al., 2019)

The benefits of workforce training in trauma-informed care are not limited to better outcomes for children and families. Organizations that adopt trauma-informed care have better treatment outcomes than non-adopters, and have lower costs as a result of staff turnover, less use of sick time, and few liability-related expenses.(Robey et al., 2021) Trauma-informed organizational culture has been associated with increased psychological wellness for behavioral health personnel and direct support personnel.(Hales & Nochajski, 2020; Keesler, 2020)

## Methods

We utilized the Project ECHO® model to deliver 12 virtual case-based interactive sessions about trauma-informed care to a diverse maternal and child health workforce from multiple care sectors and organizations in 3 California counties. Project ECHO® is an innovative real-time tele-education and tele-mentoring outreach model that was designed to address the needs of vulnerable populations by democratizing specialty knowledge to community-based providers. Using distance learning teleconferencing, Project ECHO® creates a “Hub-and-Spoke” all-teach, all-learn community that connects community providers (“Spokes”) to a multidisciplinary specialty team (“Hub”). Formal procedures were followed to establish CME and CE credit for session participants.

The ECHO series was convened for 12 bimonthly sessions over 6 months. Participants convened from geographically separate locations. The project occurred during the COVID-19 pandemic, and almost all participants joined from their homes using their own phones and computers to connect to sessions. Each 75-minute session consisted of a 15-minute didactic session, a 55-minute case-based discussion presented by a participant, and a wrap-up. The curriculum for the series followed the organization proposed by the California ACEs Aware Initiative for teaching this material: Screen-Treat-Heal. Table 1 shows the curriculum outline. [Table 1] A representative of the lead agency (the hub) facilitated the discussion, noted any systemic barriers to provision of integrated trauma-informed care, and led the discussion toward solutions to identified barriers. Rich discussions were generated between cross-sector and cross-county representatives (community spokes). The case-based presentations highlighted successful practices in place in each of the counties that could then be shared among participants. The wrap-up summarized group-generated recommendations. The last 2 sessions brought the participants into small break-out groups by sector and county to discuss barriers and solutions unique to their situation. Participant surveys were collected at enrollment (Baseline Survey), after each session (Post-Session Surveys), and at the conclusion of the series (Final Survey). All research was conducted in accord with prevailing ethical principles. This project did not meet the definition of human subjects research by the Stanford University Institutional Review Board because it was considered an educational program and therefore consent for participation and surveys was not required.

**Table 1.** Stanford ACEs Aware ECHO: Series Curriculum

| Screen | Session Goal | Case-Based Learning |
| --- | --- | --- |
| Why Screen for Adverse Childhood Experiences (ACEs)? | Demonstrate an understanding of Adverse Childhood Experiences (ACEs) | 6 year-old female placed into foster care due to parental neglect |
| How Do You Screen for ACEs? | Assess a child's ACEs using the PEARLS screen | 7 year-old female with autism |
| How Do You Respond to an ACE Screen? | Counsel and teach parents how to mitigate ACEs to reduce toxic stress | 7 year-old nonverbal child with significant medical needs and history of homelessness |
| How Do You Support Providers Who Are Doing ACEs Screenings? | Act to prevent vicarious trauma for staff who are doing PEARLS Screen | Provider needs support to manage vicarious trauma |
| Treat | Session Goal | Case-Based Learning |
| How Do You Develop a Trauma-Informed Treatment Plan? | Formulate treatment plans with knowledge of the science of ACEs | 9 year-old Latinx child; somatic complaints in response to immigration trauma |
| What Are Evidence-Based Strategies to Regulate the Stress Response? | Integrate the principles of Trauma Informed Care to treatment planning | 34 month-old male with developmental delay; family afraid to access services for fear of compromising immigration status |
| How Do You Validate Existing Strengths and Protective Factors? | Choose effective strategies for screening to reduce negative emotional responses or re-traumatization | 11 year-old male exposed to domestic violence; support needed during the pandemic |
| What Resources and Referrals Can You Recommend to Address ACEs? | Discuss ACE Screening Clinical Workflows | 3 year-old girl reunified with biological parent receives support from a Court Appointed Special Advocate |

| Heal | Session Goal | Case-Based Learning |
| --- | --- | --- |
| How Do We Cultivate Self-Compassion in Stressful Times? | Describe the key components of self-compassion and mindfulness and how you can integrate them into your professional role | 13 year-old girl in foster care who moved counties, disrupting her medical care/social-emotional/learning |
| Cultivating Cultural Compassion | Cultivate cultural compassion as an individual, as a healthcare or education professional, and as a member of a larger organization | Interactive workshop |
| How Do We Create Trauma-informed Systems of Care in Our Organizations and Communities? Part 1 | Identify the key elements of creating trauma-informed systems of care within organizations | 8 year-old girl referred for services from Child Protective Services; family relinquished parental rights; disruptions to placement and educational services due to COVID-19 pandemic |
| How Do We Create Trauma-informed Systems of Care in Our Organizations and Communities? Part 2 | Identify next steps towards creating trauma-informed systems of care in your county | 8 year-old boy with ADHD and a history of intrauterine drug exposure |

## Results

### Participation

Participants were recruited from within the lead agency’s network of community providers, and invitations were generated by email and word of mouth. A total of 100 unique participants attended the sessions, representing 54 organizations in 3 counties. The average number of participants per session was 44 (M=44.2, SD=8.41, range: 31-54). Participants each attended an average of 5 sessions out of the 12 offered (M=5.12, SD=3.74, range: 1-12). Participants included PCCs, mental health providers from Behavioral Health Service Agencies and County Family Health nursing, educators representing local school districts and Special Education Local Plan Areas, community health providers from family resource agencies, housing and parenting support organizations, and an attorney from a local medical-legal partnership. See Table 2 for number of participants and organizations by county. [Table 2] A website was developed as a communication hub for the group. (https://med.stanford.edu/aces.html) The common language for trauma-informed care developed by the state-level ACEs Aware Initiative was used for all presentations and communications.

**Table 2.** Participants Enrolled in Stanford ACEs Aware ECHO by Care Sector

| Care Sector | County |  |  |
| --- | --- | --- | --- |
|  | County 1<br>Individuals<br>(Represented<br>Organizations) | County 2<br>Individuals<br>(Represented<br>Organizations) | County 3<br>Individuals<br>(Represented<br>Organizations) |
| Healthcare Provider<br>(e.g. MD, DO, NP,<br>MSN, CNS, RN)<br>(n = 46) | 9 (7) | 20 (6) | 17 (8) |
| Behavioral Health<br>(e.g. LCSW, PhD,<br>PsyD, MFT) (n = 31) | 6 (4) | 8 (7) | 17 (7) |
| Education<br>(n = 12) | 2 (2) | 8 (3) | 2 (1) |
| Early Intervention<br>(n = 3) | 2 (1) | 1 (1) | --- |
| First 5<br>(n = 3) | 2 (1) | --- | 1 (1) |
| Family Support Agency<br>(n = 4) | 2 (2) | 1 (1) | 1 (1) |
| Legal/Justice<br>(n = 1) | 1 (1) | --- | --- |
|  | n = 24 (18) | n = 38 (18) | n = 38 (18) |

Total Participants=100 Total Organizations= 54

Participants completed Baseline Surveys generated by email, including demographic information and familiarity with and completion of a California Office of Surgeon General-recommended training. Post-session surveys were completed after each session. Each survey included questions about overall quality of the session. A voluntary Final Survey was completed electronically by a self-selected subset of the participants and requested self-reported attainment of each session’s key learning objective using a 5-point Likert scale.

All surveys were delivered electronically using Qualtrics. Data from the Baseline Survey, Post-Session Surveys and Final survey were exported into the IBM® SPSS® Version 26 software platform for analysis.

### Data Analysis

We analyzed results using chi-square for comparing whole-group proportions of dichotomous variables from Baseline to Final Surveys, and McNemar’s test and paired-sample t-tests (for dichotomous and continuous variables respectively) to measure changes within paired Baseline and Final groups.

After the sessions, participants reported that they felt more informed about ACEs and toxic stress, and better able to care for their patients using tenets of trauma-informed care. Post-Session Survey responses indicated high levels of satisfaction with the components of the sessions (didactics and case-based group discussion) and with the Zoom platform. (Table 3) [Table 3]

**Table 3.** Mean Participant-rated Score on Twelve Post-Session Surveys^a^

| Survey Question | Mean Rating Score<sup>b</sup><br>(Standard Deviation) | Range |
| --- | --- | --- |
| Overall quality of the education offered in this session | 4.78 (.09) | (4.58-4.93) |
| Session met stated objectives | 4.67 (.14) | (4.42-4.88) |
| Session enhanced my current knowledge base | 4.59 (.18) | (4.33-4.81) |
| Material presented at appropriate level | 4.64 (.13) | (4.42-4.81) |
| Instructor responsive to participants | 4.72 (.15) | (4.5-4.94) |
| Material provided useful information for work | 4.65 (.13) | (4.4-4.81) |
| I am more informed about ACEs and toxic stress, trauma-informed care, and resiliency | 4.60 (.12) | (4.4-4.75) |
| The case presentation and subsequent discussion increased my ability to better manage the care of my patients | 4.61(.14) | (4.4-4.86) |
| Group discussion made a positive impact on my educational experience | 4.60 (.17) | (4.4-4.87) |
| Questions or concerns were addressed effectively and in a timely manner | 4.68 (.14) | (4.42-4.88) |
| Zoom platform conducive to learning | 4.59 (.12) | (4.4-4.8) |
| Effectiveness of didactic speaker | 4.69 (.12) | (4.47-4.88) |

<sup>a</sup>Mean number of respondents per survey (M=19.5, SD=6.63, range: 12-29)
<sup>b</sup>5 point Likert scale 1= Strongly Disagree, 5= Strongly Agree

Active ACE screening within the participants’ organizations increased over the course of the series from 26% (28/106) pre-intervention to 45% (14/31) post-intervention, but the change was not significant. Self-reported importance of ACE screening in community care using a 5-point Likert scale remained high for participants from Baseline (M=4.69) to Final Survey (M=4.78). While not statistically significant, a key goal of the series was met when familiarity with the California ACEs Aware Initiative increased from 86% (91/106) pre-intervention to 100% (32/32) post-intervention. A significant number of ECHO participants completed the state-level recommended additional online ACEs Aware Initiative provider training curriculum during the course of the series: 25% (26/106) pre-intervention and 59% (19/32) post-intervention, X^2^(1)=11.47, p=.001 for unpaired proportions. McNemar’s test determined that the difference between pre-intervention and post-intervention proportions for paired data about completion of additional training was also statistically significant (p =.008).

In the Final Survey, participants were asked to use a 5-point Likert scale to express “how confident are you in achieving series goals *prior to* the ECHO and *now*” for key learning objectives of the series. Paired t-test analysis indicated significant change in self-reported confidence across all learning goals pre- and post-intervention. (Table 4). [Table 4]

**Table 4.**
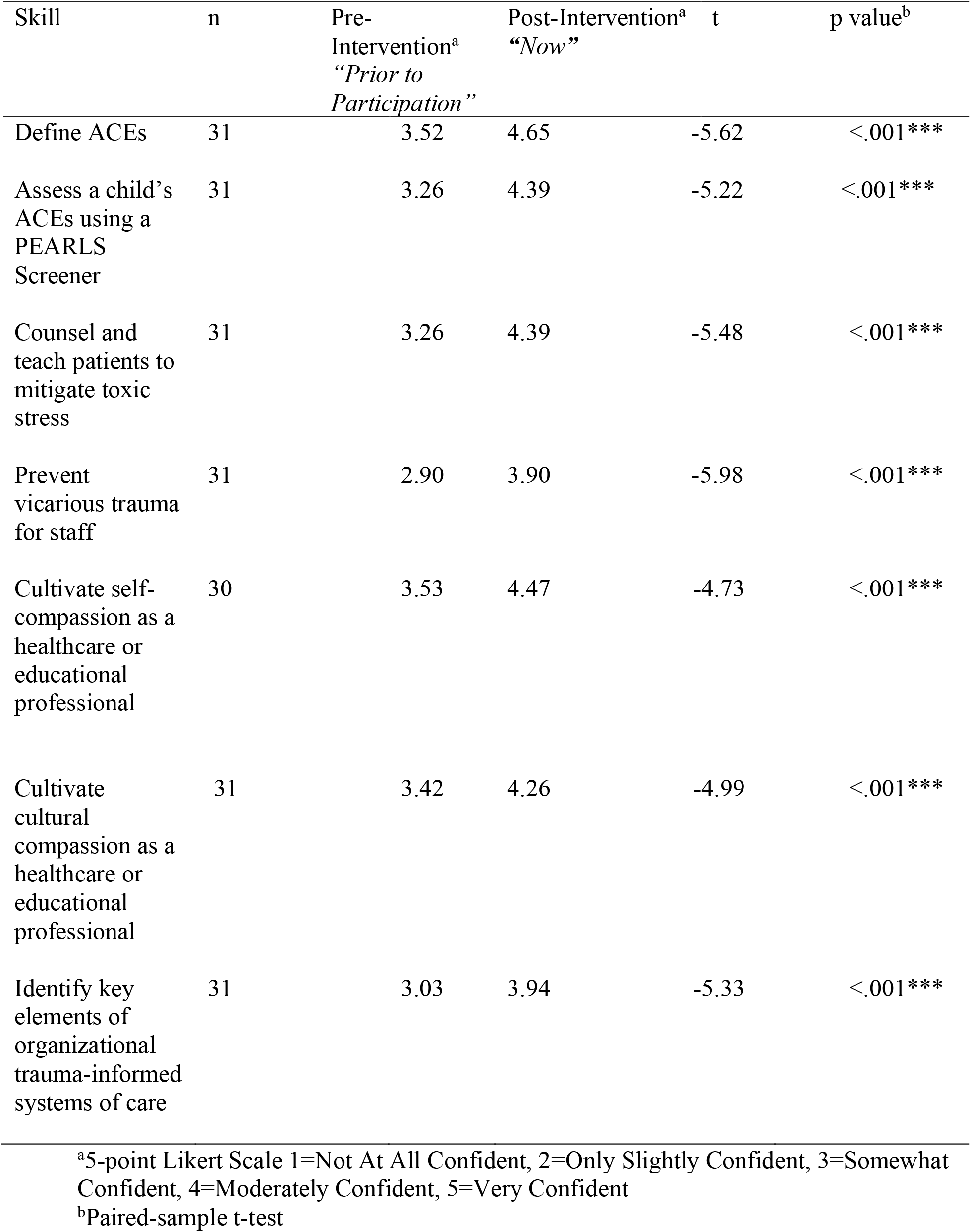
Change in Participants’ Self-Reported Confidence in Achieving Series Goals *Prior To* Participation in ECHO and *Now*

In order to determine likely changes in participant behavior after the training, the Final Survey offered several answers to the question “How will you change your clinical care based on this educational series?” Responses are recorded in Table 5. A majority of respondents reported anticipated commitment to improved cross-sector communication and referral systems. Participants were also offered several options in response to perceived barriers and opportunities to providing trauma-informed care, shown in Table 6. After participation in the ECHO series, over 2/3 of respondents reported potential to create new systems of trauma-informed care in their community. [Table 5] [Table 6]

**Table 5.** Clinical changes participants have implemented or plan to implement - More than one answer allowed (Final Survey)

| Anticipated Changes | Percent of Respondents | Number of Participants that Selected this Option |
| --- | --- | --- |
| Change in interprofessional communication or collaboration, for referrals and off-site partners | 61% | 17 |
| Change in treatment or management approach, based on ACE score and toxic stress risk assessment | 57% | 16 |
| Change in current practice for referrals or linkages to treatment and support services | 54% | 15 |
| Change in interprofessional team communication or collaboration, within team in primary care setting | 46% | 13 |
| Apply the clinical algorithm for ACEs and toxic stress identification and management to guide patient care | 29% | 8 |
| Routine screening for ACEs in adults | 11% | 3 |
| Total |  | 28 |

**Table 6.** Perceived Barriers and Opportunities to Implementing Changes – More than one answer allowed (Final Survey)

| Barriers | Percent of Respondents | Number of Participants who Selected this Option |
| --- | --- | --- |
| Lack of time during clinic workflow to implement screening | 48% | 16 |
| Lack of staff for care coordination | 39% | 13 |
| Cost of care coordination to respond to patient-reported trauma | 36% | 12 |
| Lack of time to create a clinic protocol for routine screening | 30% | 10 |
| Lack of staff to conduct ACE screening | 30% | 10 |
| Lack of referral resources if significant trauma exposure is identified | 30% | 10 |
| Potential negative staff reactions to screening | 24% | 8 |
| Potential negative patient/client reactions to screening | 15% | 5 |
| Cost of screening | 3% | 1 |
| ACEs not clinically relevant | 0% | 0 |
| Total |  | 33 |

| Opportunities | Percent of Respondents | Number of Participants who Selected this Option |
| --- | --- | --- |
| Improved patient care | 88% | 29 |
| Potential to create new systems of trauma-informed care in my community | 82% | 27 |
| Financial benefits for clinics with insurance reimbursement | 33% | 11 |
| Total |  | 33 |

## Discussion

The Stanford ACEs Aware ECHO effectively increased participants’ knowledge and confidence to identify ACEs and provide TIC within their communities. This virtual 12 session workforce development series introduced curricular content created by the California ACEs Aware Initiative and was associated with a significant increase in uptake of an additional key training offered by the ACEs Aware Initiative to its target maternal and child workforce. The majority of learners rated the Stanford ACEs Aware ECHO sessions to be effective from a technical and learning standpoint. During the sessions, high levels of cross-county and cross-sector dialogue occurred, as participants discussed real de-identified cases. After completing the series, participants across maternal child health care sectors self-reported significant increases in confidence to use best-practice skills in their work. Delivered virtually in the midst of the COVID-19 pandemic to participants across geographic areas and care sectors, this project provides promising data that supports the efficacy of virtual interactive teaching models in training a maternal-child workforce that can effectively deliver best practice trauma-informed care across systems.

The curriculum development and delivery process were vastly simplified by the use of the Project ECHO® model. Use of this model has been well-established in similar situations, and provided a framework upon which to lay content. We anticipate that the videoconferencing delivery model can be easily generalized for different content and geographic delivery areas. ECHO uses a “learning by doing” and “guided practice” model, which allows knowledge dissemination and standardization of care by promoting best practices to geographically isolated participants.

Previous studies have shown that Project ECHO® dramatically improves both capacity and access to specialty care for rural and underserved populations. As participants gain increasing independence and their skills and self-efficacy grow through involvement in an ongoing community-based learning program, participant career satisfaction has been shown to improve. (Arora et al., 2017) Similar to published research on ECHOs, we found that after participation in the educational series, maternal and child health professionals reported increased comfort in treating a clinical condition (toxic stress) they had previously experienced as challenging.(Arora et al., 2011) Like others before, we found that the ECHO model was an effective way to bring evidence-based best practice care to multiple professionals in different care sectors.(Nakamura et al., 2019)

A challenge of running an ECHO is finding committed and representative partners. Asking busy clinicians and agency representatives to allocate time to attend sessions is not easy. In order to create a true cross-sector collaboration also requires knowledge of who needs to be at the table to provide cross sector care. We were able to overcome these challenges because of deep professional ties within care communities and word of mouth promotion of the project from key partners with the target care sectors and counties. Free CME and CEU credits and strong interest in the curricular topics prompted participants to attend the sessions without financial support. Holding the sessions over the noon hour allowed participants to attend during their lunch break. Future projects might consider inviting initial partners who have deep professional community ties to recruit local participants from within target maternal and child health workforce groups and care sectors. Additionally, if feasible, small monetary stipends may allow organizations to dedicate an individual for the proposed number of hours to attend the educational series. A limitation of the project is that although we surveyed participants about “anticipated change” we did not have behavioral data to determine the practice change effects of the educational experience. Ideally, we would gather data from the clinicians’ electronic health record, interagency partners and/or from patient families about changes in provider practice after participation.

In summary, this project demonstrated the feasibility of delivering a curriculum about TIC to a virtual community of cross-sector, multi-agency maternal child health workforce professionals during the COVID pandemic using the ECHO model. Participation in the series was associated with increased uptake of a vital recommended training developed by the California Office of the Surgeon General and Department of Healthcare Services, and increased participants’ self-reported knowledge and confidence to provide evidence-based best practice trauma-informed care within their communities.

## Data Availability

Data will be made available by request to corresponding author.

## Acknowledgements

This project was supported by funding by the California Office of the Surgeon General and Department of Health Care Services’ ACEs Aware Initiative Provider Engagement (Network of Care) grant.

The authors extend their gratitude to Lettie McGuire, EdM, Anne DeBattista PhD, MSN, and Emily Whitgob MD, MEd for their invaluable contributions to this project.

